# The influence of sex, gender, age, and ethnicity on psychosocial factors and substance use throughout phases of the COVID-19 pandemic

**DOI:** 10.1101/2021.06.08.21258572

**Authors:** Lori A. Brotto, Kyle Chankasingh, Alexandra Baaske, Arianne Albert, Amy Booth, Angela Kaida, Laurie W. Smith, Sarai Racey, Anna Gottschlich, Melanie C.M. Murray, Manish Sadarangani, Gina S. Ogilvie, Liisa Galea

## Abstract

The SARS-CoV-2 (COVID-19) pandemic has had profound physical and mental health effects on populations around the world. Limited empirical research has used a gender-based lens to evaluate the mental health impacts of the pandemic, overlooking the impact of public health measures on marginalized groups, such as women, and the gender diverse community. This study used a gender-based analysis to determine the prevalence of psychosocial symptoms and substance use by age, ethnicity, income, rurality, education level, Indigenous status, and sexual orientation.

Participants in the study were recruited from previously established cohorts as a part of the COVID-19 Rapid Evidence Study of a Provincial Population-Based Cohort for Gender and Sex (RESPPONSE) study. Those who agreed to participate were asked to self-report symptoms of depression, anxiety, pandemic stress, loneliness, alcohol use, and cannabis use across five phases of the pandemic as well as retrospectively before the pandemic.

For all psychosocial outcomes, there was a significant effect of time with all five phases of the pandemic being associated with more psychosocial symptoms relative to pre-COVID levels (*p <* .0001). Gender was significantly associated with all outcomes (*p <* .0001) with men exhibiting lower scores (i.e., less symptoms) than women and gender diverse participants, and women exhibiting lower scores than the gender diverse group. Other significant predictors were age (younger populations experiencing more symptoms, *p <* .0001), ethnicity (Chinese/Taiwanese individuals experiencing less symptoms, *p* = .005), and Indigenous status (Indigenous individuals experiencing more symptoms, *p* < .0001). Alcohol use and cannabis use increased relative to pre-pandemic levels, and women reported a greater increase in cannabis use than men (*p* < .0001). Our findings highlight the need for policy makers and leaders to proactively consider gender when tailoring public health measures for future pandemics.

## Introduction

In the first eighteen months of the SARS-CoV-2 (COVID-19) pandemic, there have been over 150 million cases and over 3 million deaths attributable to the upper respiratory virus [1]. More specifically, Canada has reached a stark milestone of one million cases and over 20,000 deaths in a little over a year (May 2021) [2]. Although the physical health effects of the virus tend to dominate the literature and the media, it is well established that outbreaks, including the current pandemic, have significant impacts on the mental health of those involved. For example, healthcare workers and patients affected by previous outbreaks such as SARS-CoV-1 [3], H1N1 influenza [4], and Ebola [5] have poorer psychosocial outcomes during the onset of societal alarm.

Public health measures put in place due to the COVID-19 pandemic have had a negative impact on the mental health of peoples worldwide [6, 7]. Levels of depression [8], anxiety [8], loneliness [9], alcohol use [10, 11], and cannabis use [12] have all increased relative to pre-pandemic levels. Additionally, there is mounting evidence highlighting the secondary effects of public health measures on specific populations during the pandemic [13]. For example, younger populations [14] and those of lower income [14] have experienced disproportionate psychosocial outcomes because of the COVID-19 pandemic.

There is a growing realization that an intersectional lens needs to be applied to COVID-19 research, not only regarding biomedical outcomes, but for psychosocial outcomes as well [15]. Across North America, more efforts are being directed towards sex and gender research. Sex is defined as birth assignment and is usually established by genital anatomy at birth with female, male, and intersex as typical response options in queries about sex. Gender identity is defined as one’s personal feelings about being a woman, man, transgender, gender-diverse individual, or another expression of gender that does not align with that person’s birth assigned sex. When sex is considered in the context of psychosocial issues, it is well established that females are more likely to present symptoms of depression and anxiety outside of the pandemic [16] and have faced greater job losses than males during the COVID-19 pandemic [17]. Thus, it is not surprising that studies to date have found that females have reported more anxiety, depressive symptoms and post-traumatic stress symptoms relative to males during the COVID-19 pandemic [14,18–20]. Age also plays a large role in sex differences in the risk for neuropsychiatric disorders [21], but thus far the interaction between age and sex has received little attention with regards to how age may interact with sex and psychosocial outcomes throughout COVID-19.

In addition to the paucity of sex-based analyses, studies examining psychosocial outcomes from the standpoint of participants’ self-identified gender are few. Most of the research on gender and the COVID-19 pandemic have compared responses between women and men, while ignoring the experiences of individuals who experience gender on a spectrum, beyond the binary classification of man and woman. A recent cross-sectional survey by Hawke et al. [22] found that despite no clear significant differences in mental health between cisgender, transgender and gender diverse youth before the pandemic, those identifying as gender diverse were two times more likely to report experiencing mental health challenges relative to the cisgender group during COVID-19. These findings were associated with an unmet need for mental health and substance use services. However, this study was limited to those aged 14-28, thereby reducing the generalizability of the findings to the larger population. To our knowledge, no studies have taken a gender-based approach to assess the prevalence of mental health sequelae of COVID-19 pandemic control measures across a general population sample. Given the current data as well as previous findings on the poor mental health outcomes of gender diverse individuals [23], focused empirical attention on this population is critical.

Other social determinants of health (SDoH), including education, ethnicity, and income, impact physical and mental health outcomes [24] and have shaped the risk and consequences of COVID-19 in communities across North America [25]. Additionally, minority stress theory has posited that those who are part of more than one marginalized societal group may experience even greater health disparities [26]. Given this, it is crucial to not only explore the impact of the SDoH on health outcomes, but also how these factors interact with each other and with gender to influence mental health. A year and a half into the pandemic, gaps in research exploring these intersections remain. Our study presents a unique opportunity to explore the sex and gendered effects associated with the unintended consequences of the COVID-19 pandemic.

Many governments have tailored public health interventions throughout the pandemic based on infection incidence and hospitalization rates, resulting in a series of lockdowns (and prescribed regulations), followed by periods of relaxed restrictions, which have generated defined “phases” of the pandemic. While it is now widely known that lockdowns impact mental health [27], little research has explored mental health across these different “phases” of the pandemic.

We examined the prevalence of psychosocial outcomes by sex, gender, age, ethnicity, income, education, rurality, and sexual orientation as well as how these characteristics may interact with each other to impact the mental health of a large cohort of the general population in British Columbia (BC) across different phases of the pandemic. First, we examined the prevalence of depression, anxiety, stress, and loneliness based on previous findings reporting that these outcomes have been impacted by the pandemic, and secondly, we measured the associations of these psychosocial variables with increased alcohol and cannabis consumption. We predicted that these four psychosocial domains would be more symptomatic in women and gender diverse individuals than in men and that this would be influenced by age, ethnicity, and income. We also hypothesized that during phases of increased social restriction psychosocial symptoms would increase.

## Materials and Methods

### Participant Recruitment and Study Design

Participants, aged 25-69 years, were invited to participate in this study from previously established cohorts from the XXXXX (information blinded for peer review), representing both general and priority populations of BC who had consented to being contacted for future research [28]. Participants were stratified into nine five-year age strata, and using a SARS-CoV-2 population seroprevalence of 2% (±1, 95% CI), the target recruitment for each stratum was 750. The seroprevalence statistic was used to target recruitment for analyses in a separate manuscript.

Those identified as potentially eligible from the established cohorts (*Index Participants)* were sent an email invitation to participate via an online survey. To improve the representativeness of the study sample, Index Participants were asked to pass the invitation on to one household member who identified as a different gender as the respondent *(Household Participants)*. All potential participants were sent up to three email reminders to participate in the study. The inclusion criteria were: current residents of BC, aged 25-69, any gender, and able and willing to fill out the online survey in English. Ethics approval was obtained from the XXXXXXX Research Ethics board (information blinded for peer review). Survey responses were collected anonymously, with the exception of three-digit postal codes, which were used to determine rurality for analyses.

After two months of data collection from existing research cohorts, recruitment was expanded to include participants who were not from previous cohorts, as well as public recruitment through the REACH BC platform, social media (i.e., Facebook, Twitter and Instagram), posts on the XXXX website and engagement of community groups and other stakeholders to facilitate recruitment. All respondents in the study were invited to enter a draw to win a $100 e-gift card for completing the survey. Recruitment was continued until a target of n = 750 was reached for each of the nine age-based strata, with the exception of the 25-29 year age group. Recruitment was open from August 20, 2020 – March 1, 2021.

### Survey Design and Measures

The survey was tested for face validity, pilot tested, and a final version was designed using REDCap (Research Electronic Data Capture) [29]. While the survey consisted of multiple modules, this study focuses solely on the outcomes from the psychosocial module, which included questions about mental health outcomes such as depression, anxiety, stress, loneliness, alcohol use, and cannabis use.

Demographic information was collected from all respondents including age, sex, gender, sexual orientation, ethnicity, Indigenous status, income, education level, if the participant was currently a student and rurality by postal code. Sex referred specifically to the sex assigned at birth and included the option of male, female or intersex. Gender referred to the respondent’s current gender identity and included the options man, woman, or another option grouping non-binary, transgender, GenderQueer, agender or any other similar identity together. Sexual orientation options included asexual, bisexual, demisexual, gay/lesbian, heterosexual, or pansexual. Participants were given the option to identify as the following ethnicities: White, Chinese/Taiwanese, Black (African, Caribbean, or Other), South Asian (e.g., Indian, Bangladeshi, Pakistani, Punjabi, and Sri Lankan), and several other ethnicities who were analyzed an “Other” category. Indigenous status was assessed separately from ethnicity. Self-reporting of Indigenous status provided participants the option to identify as First Nation, Metis, Inuit, non-status First Nations, other Indigenous or not Indigenous, and they were then asked about Two Spirit identity. Rurality was determined based on three-digit postal codes and were classified into one of the follow categories: census metropolitan area, strong metropolitan influence zone, moderate metropolitan zone, or weak to no metropolitan influence zone.

Although the survey was completed at only one cross-sectional time point, some of the questions asked participants to refer to different periods of time: pre-pandemic (before March 2020) as well as across five different phases of the pandemic in BC. In the first version of the survey, Phase 1 lasted from mid-March 2020 to mid-May 2020, Phase 2 lasted from mid-May 2020 to mid-June 2020, and Phase 3 lasted from mid-June 2020 until the end of November 2020. At the end of November 2020, the survey was amended to include Phases 4 and 5, as well as modified dates for Phases 2 and 3 (mid-May to end of August 2020; denoted by Phase 2/3_2). Phase 4 lasted from September 2020 to the end of October 2020 and Phase 5 lasted from November 2020 to the date our survey closed (March 1, 2021). We have included a Supplemental Table that explains the public health recommendations in more detail, through every phase of the pandemic in BC.

#### Depression

Depression was measured across the phases of the pandemic using the Patient Health Questionnaire (PHQ-9). The PHQ-9 questionnaire was used to measure self-reported symptoms of depression on a Likert scale from zero (not at all) to three (nearly everyday). Scores for this questionnaire range from 0-27 with a score of 0-4 indicating minimal depression, 5-14 indicating mild to moderate depression and 15-27 indicating moderately severe to severe depression [30]. The PHQ-9 has been validated across age and gender, as well as among diverse populations [30, 31]. Internal consistency across data collection and Cronbach’s alpha for the PHQ-9 in the current sample was very good at α = 0.848.

#### Anxiety

Anxiety was measured across the phases of the pandemic using the Generalized Anxiety Disorder questionnaire (GAD-7). The GAD-7 was provided to respondents to self-report feelings of anxiety on a Likert scale from zero (not at all) to three (nearly everyday). Scores for this questionnaire range from 0-21 with scores above 10 indicating a clinical diagnosis for anxiety [32]. The GAD-7 has been validated in the general population and is frequently used in primary care settings to screen for anxiety symptoms[33]. Internal consistency across data collection and Cronbach’s alpha for the GAD-7 questionnaire in the current sample was very good at α = 0.889.

#### Pandemic Stress

General pandemic stress was measured across the phases of the pandemic using the CoRonavIruS Health Impact Survey (CRISIS) V0.3. This survey was developed and validated early in the COVID-19 pandemic to provide a general measure of mental distress and resilience [34]. Participants were asked to self-report feelings of stress on a Likert scale from one (not at all) to five (extremely). Scores for this questionnaire range from 10-50 with higher scores indicating greater COVID-related stress. Internal consistency across data collection and Cronbach’s alpha for CRISIS in the current sample was very good at α **=** 0.882.

#### Loneliness

Loneliness was also measured across the phases of the pandemic where respondents were asked to self-report feelings of loneliness on a Likert scale from one (not lonely at all) to five (extremely lonely). Previous studies have found loneliness to be positively correlated with both PHQ-9 and GAD-7 scores [35] and to be a significant predictor of suicide [36, 37].

#### Alcohol Use

Change in alcohol use was asked for all post-COVID time points (i.e., Has your consumption of alcohol changed since March 2020?). Change in alcohol use was defined as “none” (which included no alcohol use, decreased alcohol use, and same alcohol use) vs. increased alcohol use. Therefore, a single, non-time-varying alcohol change variable was created and used in a longitudinal model with time.

#### Cannabis Use

Change in cannabis use was asked for all post-COVID time points (Has your consumption of cannabis changed since March 2020?). As with alcohol, change in cannabis use was defined as “none” (which included no cannabis use, decreased cannabis use, and same cannabis use) vs. increased cannabis use. A single, non-time-varying cannabis change variable was created and used in a longitudinal model with time.

### Statistical Analyses

Analyses were carried out using R v.4.0.3. Longitudinal analyses of psychosocial outcomes across the pandemic control phases were conducted using mixed-effects linear regression models with individual and household IDs as random effects. This allows for correlations among individuals in the same household, and separately, correlations over time among responses within the same individual. We included pairwise interactions to assess non-additive effects between pandemic phase, sex, gender, ethnicity, sexual orientation, income, and Indigenous status. Significance was assessed using likelihood-ratio tests, and interactions were removed from the models if non-significant at *p* < .05. Post-hoc pairwise tests were conducted to further explore main or interaction effects with Bonferroni correction for multiple tests.

To explore associations between increase in alcohol and cannabis use with sex/gender and other demographic variables we used mixed-effects logistic regressions with household ID as a random effect. We also examined increase in alcohol and cannabis use and psychosocial outcomes across the phases as described above. Interactions and post-hoc tests were handled as above. Missing data were excluded from analyses.

## Results

### Survey Participants

Between August 2020 and March 2021, 16,056 survey invites were emailed to prospective Index Participants and 1,872 participants were recruited from the public, for a total of 17,928 prospective participants. Of these participants, a total of 5,415 responded to the invitation to participate in the study and met the analysis inclusion criteria (Fig 1). Of these participants, 1,434 forwarded the survey invitation to a household member of a different sex or gender and we received 661 participants via this method. The present analyses includes the 6,076 Index and Household participants who completed psychosocial measures of anxiety, depression, stress, and loneliness.

**Fig 1.**
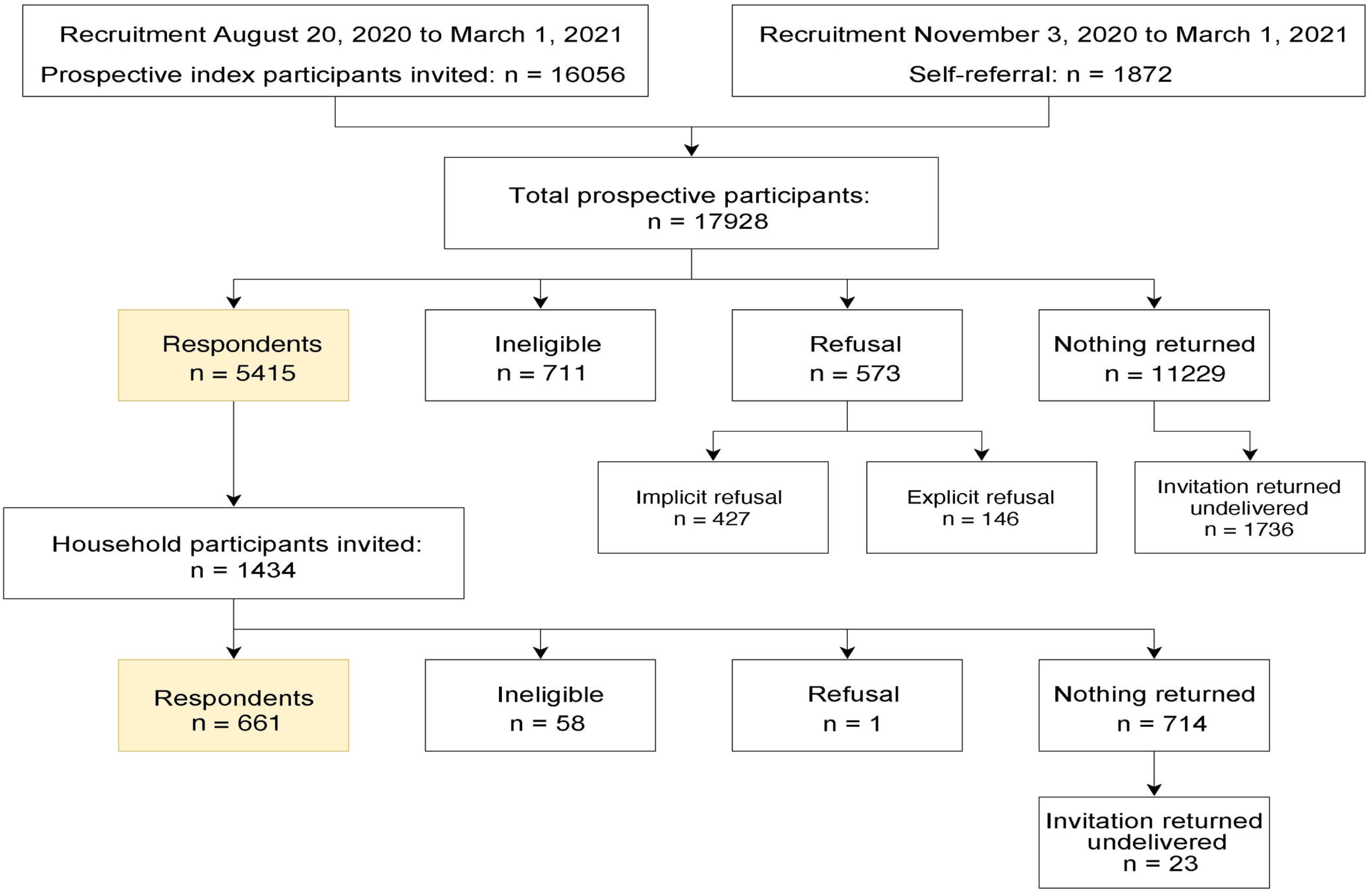
A flow diagram of prospective participants and respondents to the study.

### Demographic Characteristics of Participants

A total of 6,426 individuals responded to the question about sex; there were n = 820 males (12.7%) and n = 5,606 females (87.1%). A total of 6,076 responded to the question about gender; including men (n = 750; 12.3%), women (n = 5,254; 86.4%), and gender diverse (n = 72; 1.2%) individuals. Table 1 presents the demographic characteristics of the sample by gender.

**Table 1.** Demographic information of survey respondents. M refers to the mean age and *(SD)* standard deviation of participants.

| **Total**<br>**N = 6,076** | **Women**<br>**n = 5,254** | **Men**<br>**n = 750** | **Gender Diverse**<br>**n = 72** |
| --- | --- | --- | --- |
| **Age *M* (*SD*)** | 48.5 ($\pm 12.0$) | 48.5 ($\pm 12.2$) | 42.4 ($\pm 11.6$) |
| 25-29 | 348 (6.6%) | 68 (9.1%) | 8 (11.1%) |
| 30-39 | 1,075 (20.5%) | 133 (17.7%) | 29 (40.3%) |
| 40-49 | 1,291 (24.6%) | 164 (21.9%) | 14 (19.4%) |
| 50-59 | 1,333 (25.4%) | 212 (28.3%) | 11 (15.3%) |
| 60-69 | 1,207 (23.0%) | 173 (23.1%) | 10 (13.9%) |
| **Sexual Orientation**<br>**(%)** |  |  |  |
| Heterosexual | 4,480 (85.3%) | 674 (89.9%) | 7 (9.7%) |
| Non-Heterosexual | 757 (14.4%) | 74 (9.9%) | 65 (90.3%) |
| **Ethnicity (%)** |  |  |  |
| White | 4,265 (81.2%) | 609 (81.2%) | 44 (61.1%) |
| Black | 28 (0.5%) | 6 (0.8%) | 1 (1.4%) |
| Chinese/Taiwanese | 311 (5.9%) | 40 (5.3%) | 2 (2.8%) |
| South Asian | 123 (2.3%) | 9 (1.2%) | 1 (1.4%) |
| Other ethnicity | 504 (9.6%) | 81 (10.8%) | 20 (27.8%) |
| **Indigenous Status**<br>**(%)** |  |  |  |
| Indigenous | 166 (3.2%) | 27 (3.6%) | 11 (15.3%) |
| Not Indigenous | 4,830 (91.9%) | 696 (92.8%) | 59 (81.9%) |
| **Current Student (%)** |  |  |  |
| Yes | 311 (5.9%) | 37 (4.9%) | 14 (19.4%) |
| No | 4,939 (94.0%) | 711 (94.8%) | 58 (80.6%) |
| **Level of education (%)** |  |  |  |
| High school or less | 641 (12.2%) | 92 (12.3%) | 13 (18.1%) |
| More than high school | 4,605 (87.6%) | 658 (87.7%) | 58 (80.6%) |
| **Current Household Income (%)** |  |  |  |
| <\$10,000 – \$20,000 | 128 (2.4%) | 14 (1.9%) | 7 (9.7%) |
| \$20,000 – \$40,000 | 269 (5.1%) | 23 (3.1%) | 14 (19.4%) |
| \$40,000 – \$60,000 | 458 (8.7%) | 32 (4.3%) | 10 (13.9%) |
| \$60,000 – \$80,000 | 488 (9.3%) | 56 (7.5%) | 5 (6.9%) |
| \$80,000 – \$100,000 | 642 (12.2%) | 67 (8.9%) | 8 (11.1%) |
| \$100,000 – \$150,000 | 1,104 (21.0%) | 156 (20.8%) | 14 (19.4%) |
| >\$150,000 | 1,273 (24.2%) | 304 (40.5%) | 6 (8.3%) |
| **Rurality** |  |  |  |
| Census metropolitan area | 4,957 (94.3%) | 716 (95.5%) | 70 (97.3%) |
| Strong metropolitan influence zone | 79 (1.4%) | 7 (0.9%) | 0 (0.0%) |
| Moderate metropolitan influence zone | 116 (2.0%) | 8 (1.1%) | 1 (1.4%) |
| Weak to No metropolitan influence zone | 46 (0.8%) | 5 (0.67%) | 0 (0.0%) |

*Note:* Values do not add up to 100% due to missing data.

### Effect of Pandemic Phase, Age, Ethnicity and Gender and Sex on Psychosocial Outcomes

Controlling for household income, we found no significant interactions between age and gender, age and sex, age and ethnicity or rurality on any of the psychosocial measures. For all psychosocial outcomes, there was a significant relationship with pandemic phase (all *p* < .0001, Table 2), with the greatest increases in psychosocial symptoms in Phase 1 compared to pre-COVID. The scores in all subsequent phases remained significantly higher (i.e., more symptoms) than in the pre-COVID phase across all outcomes (Fig 2A-D, Table 2). Gender was significantly associated with all outcomes (all *p* < .0001, Fig 2A-D, Table 2), and pairwise comparisons showed that men had lower scores than both women and gender-diverse participants, while women had lower scores than the gender-diverse participants. Age was significantly negatively associated with all the outcomes, with older participants having lower scores on average (i.e., fewer psychosocial symptoms) (*p* < .0001, Table 2). Finally, there was a significant relationship between ethnicity and all outcomes (GAD-7 and PHQ-9 *p* < .0001, CRISIS and Loneliness *p* = .005, Table 2), with scores lower in Chinese/Taiwanese participants compared to the White, South Asian, and Other ethnicity participants. When sex was included in the model in place of gender, there were no differences to the findings, indicting the overlap in our participants self-reported sex and gender. Given our intention to explore outcomes separately for gender-diverse individuals, all subsequent analyses were done by gender (not sex).

**Fig 2.**
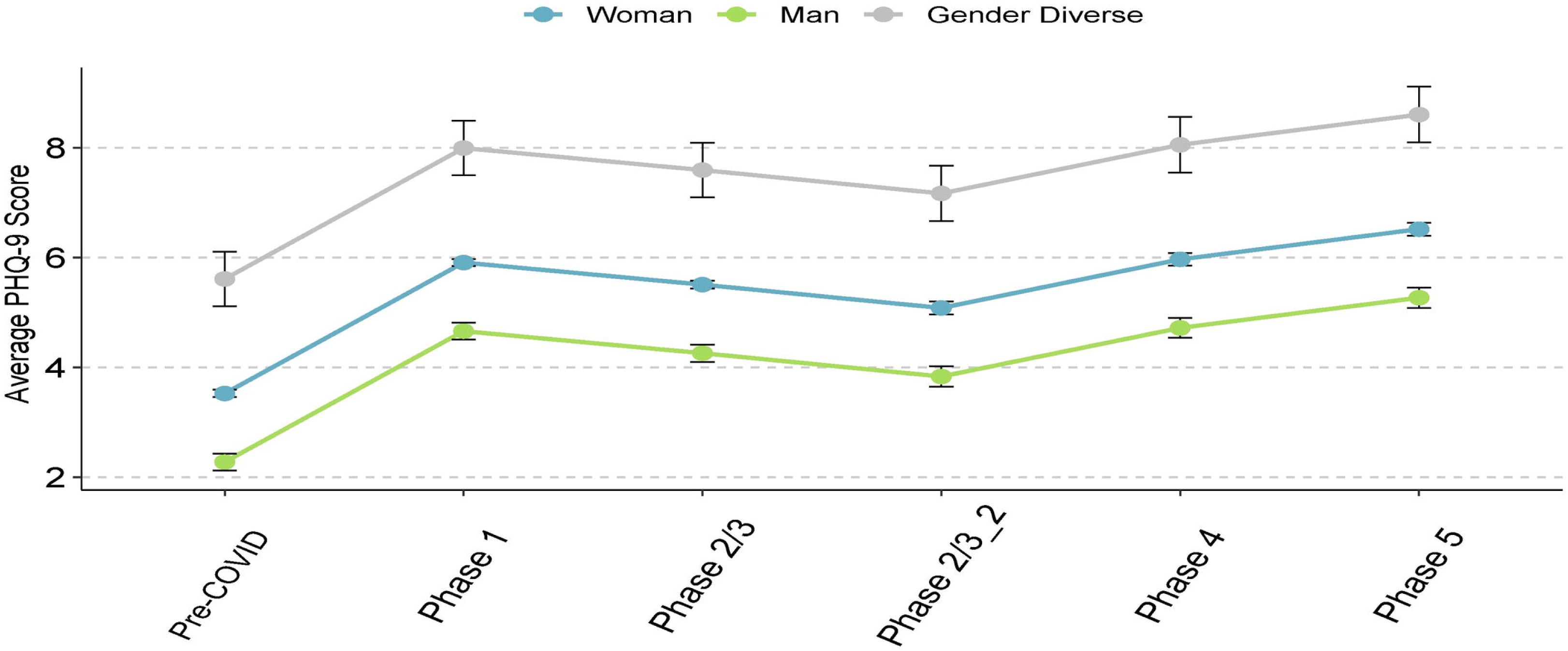
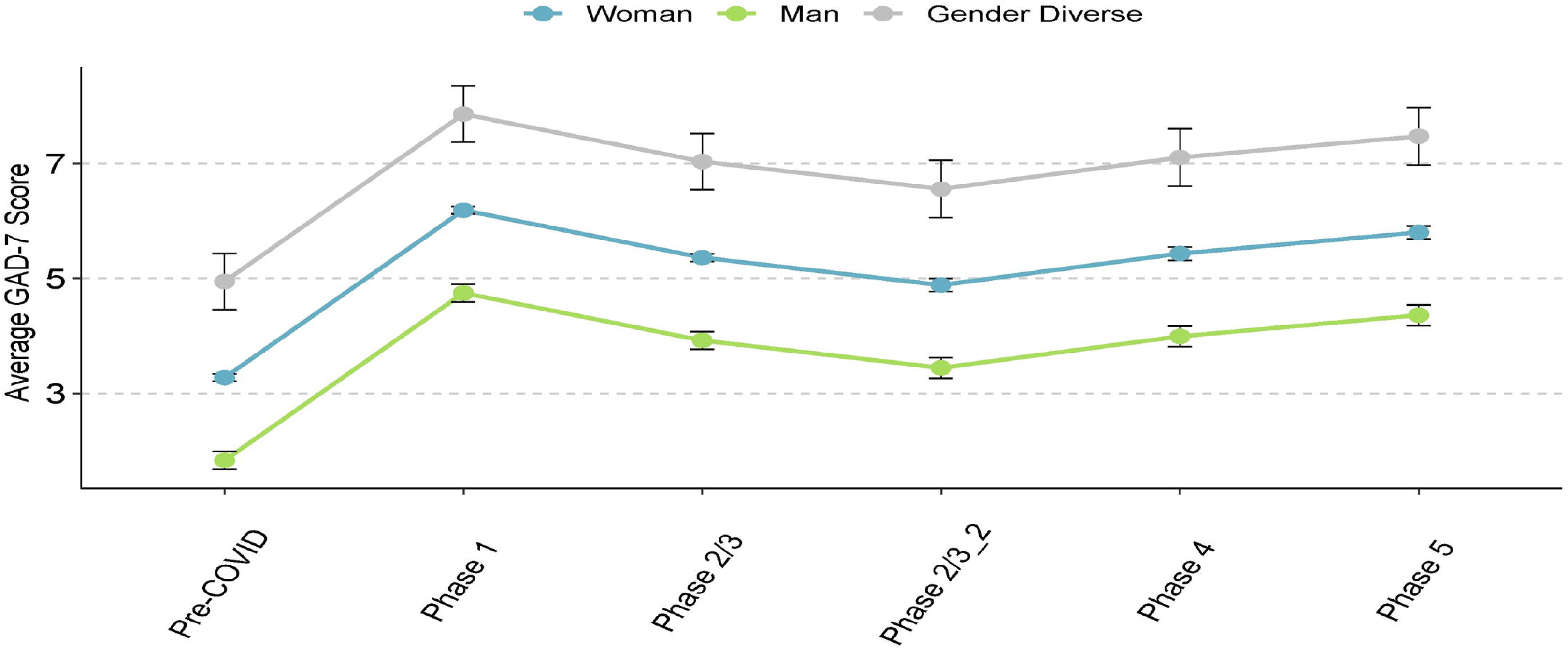
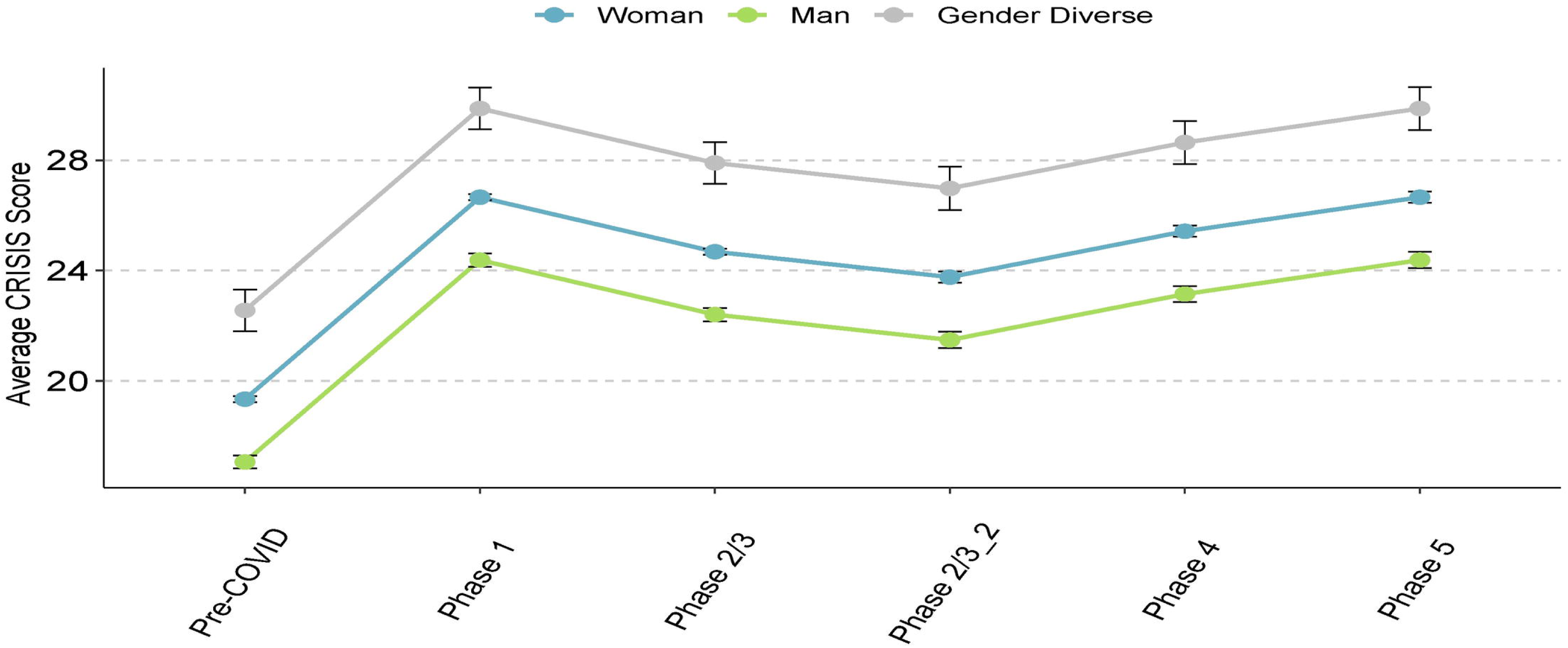
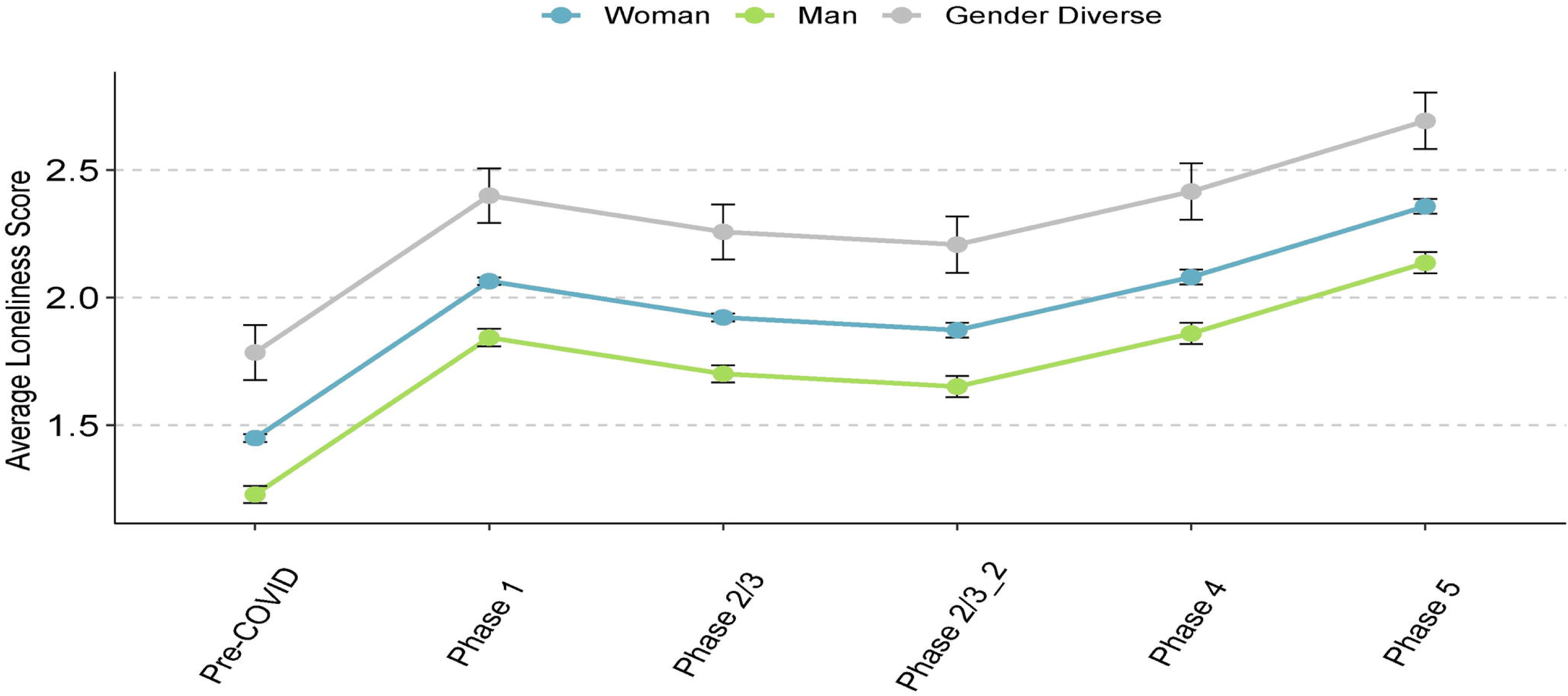
Psychosocial outcome scores across phases of the pandemic. Data points refer to mean scores of the given psychosocial measure, error bars refer to the standard error. Pre-COVID: prior to mid-March 2020; Phase 1: mid-March 2020 to mid-May 2020; Phase: 2/3: mid-May 2020 to November 2020; Phase 2/3_2: mid-May 2020 to August 2020; Phase 4: September 2020 to October 2020; Phase 5: November 2020 to March 1, 2021. (A) PHQ-9 is the Patient Health Questionnaire, a measure of depression. (B) GAD-7 is the Generalized Anxiety Disorder score, a measure of anxiety. (C) CRISIS refers to the CoRonavIruS Health Impact Survey, a measure of pandemic stress. (D) Loneliness was measured using a single data point over the five phases of the pandemic.

**Table 2.**
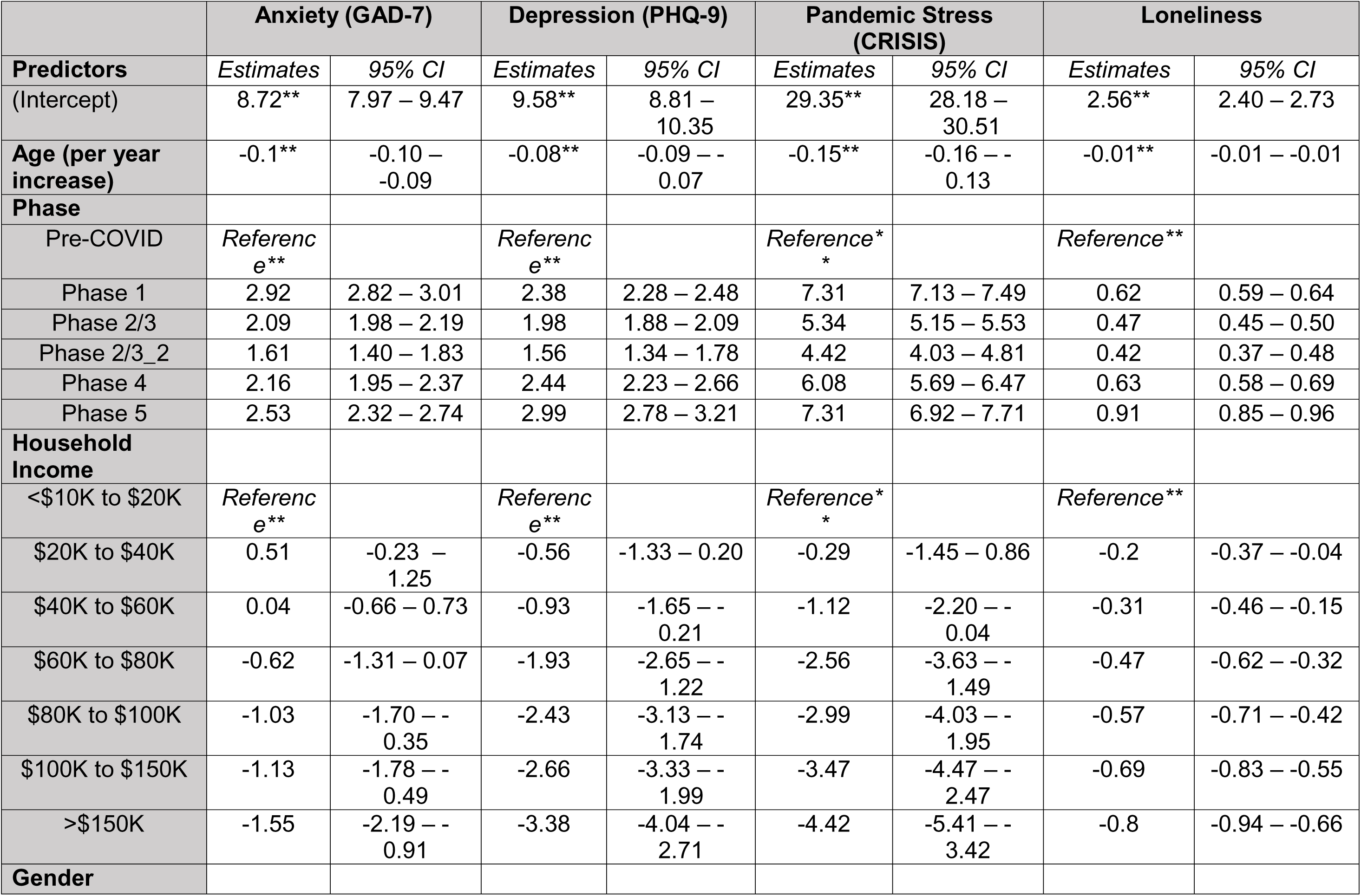
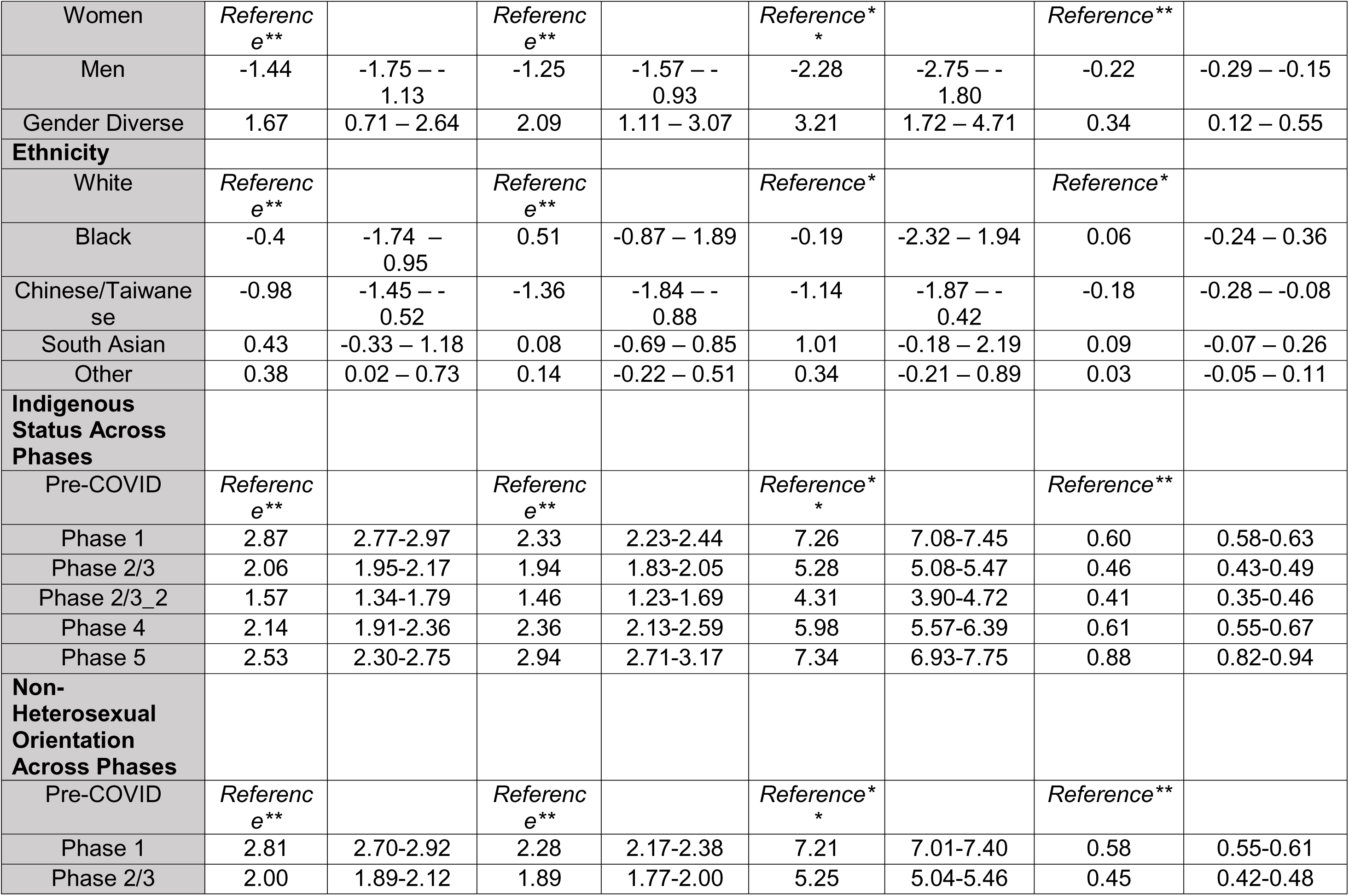
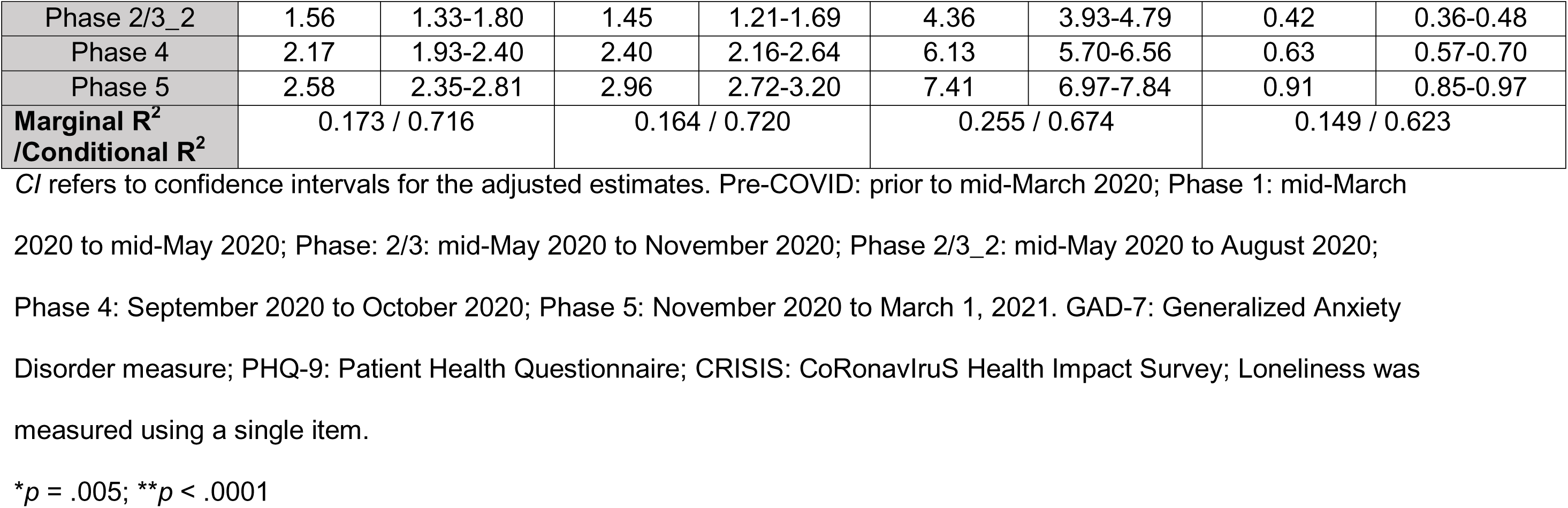
Impact of sociodemographic factors and pandemic phase on psychosocial outcomes.

### Psychosocial Outcomes by Indigenous Status

Controlling for household income, there was no significant interaction between Indigenous status and age or gender. There was a significant interaction between Indigenous status and time for all four psychosocial outcomes (*p* < .0001, Table 2) and follow-up post-hoc pairwise tests suggest that at all time points except pre-COVID, those who identified as Indigenous had significantly higher GAD-7, PHQ-9, CRISIS, and loneliness scores (i.e., more symptoms) than those who did not identify as Indigenous.

### Psychosocial Outcomes by Sexual Orientation

Across all outcomes, the non-heterosexual group (which included asexual, bisexual, demisexual, gay/lesbian, pansexual, and other) had significantly more psychosocial symptoms than the heterosexual group for all phases, and the magnitude of the difference between the groups was largest in Phase 1 of the pandemic.

### Associations between Psychosocial Outcomes and Alcohol by Gender

A total of 23.3% of the sample reported an increase in alcohol use. Increased alcohol use was negatively associated with age (*p* < .001, Table 3), with older participants having lower odds of increased alcohol use. There was no significant difference among genders in the odds of increased alcohol use, but there was a trend of increasing odds as household income increased. Additionally, those residing in census metropolitan areas were found to have increased their alcohol use relative to those outside of these dense urban areas (*p* = .03, Table 3).

**Table 3.** Changes in alcohol and cannabis use across sociodemographic factors.

| **Predictors** | **Change in Alcohol Use** |  | **Change in Cannabis Use** |  |
| --- | --- | --- | --- | --- |
|  | *OR* | *95% CI* | *OR* | *95% CI* |
| **Age** | 0.98** | 0.97 – 0.98 | 0.97** | 0.96 – 0.99 |
| **Household income** |  |  |  |  |
| <\$10K to \$20K | *Reference\** | | *Reference* | |
| \$20K to \$40K | 1.16 | 0.64 – 2.11 | 1.5 | 0.67 – 3.37 |
| \$40K to \$60K | 1.19 | 0.68 – 2.08 | 1.32 | 0.60 – 2.91 |
| \$60K to \$80K | 1.13 | 0.66 – 1.95 | 1.67 | 0.76 – 3.70 |
| \$80K to \$100K | 1.28 | 0.75 – 2.18 | 1.39 | 0.64 – 3.03 |
| \$100K to \$150K | 1.45 | 0.86 – 2.43 | 1.24 | 0.59 – 2.61 |
| >\$150K | 1.62 | 0.97 – 2.70 | 1.17 | 0.55 – 2.48 |
| **Rurality** |  |  |  |  |
| Census metropolitan area | *Reference\** |  | *Reference* |  |
| Strong metropolitan influence zone | 0.93 | 0.48 – 1.82 | 0.61 | 0.19 – 2.00 |
| Moderate metropolitan influence zone | 1.42 | 0.85 – 2.36 | 0.84 | 0.37 – 1.91 |
| Weak to No metropolitan influence zone | 0.24 | 0.07 – 0.81 | 2.15 | 0.48 – 9.61 |
| **Gender** |  |  |  |  |
| Women | *Reference* |  | *Reference\** |  |
| Men | 0.86 | 0.69 – 1.06 | 0.66 | 0.43 – 1.02 |
| Gender Diverse | 0.72 | 0.32 – 1.61 | 0.84 | 0.34 – 2.11 |
| **Ethnicity** |  |  |  |  |
| White | *Reference* |  | *Reference* |  |
| Black | 1.17 | 0.49 – 2.82 | 2.42 | 0.66 – 8.89 |
| Chinese/Taiwanese | 0.68 | 0.45 – 1.02 | 0.57 | 0.24 – 1.33 |
| South Asian | 0.89 | 0.52 – 1.53 | 0.38 | 0.10 – 1.40 |
| Other | 0.94 | 0.72 – 1.22 | 1.56 | 1.02 – 2.37 |

*OR* refers to the odds ratio, *CI* refers to the confidence intervals for the *OR*.
\* $p = .03$ ; \*\* $p < .001$

Controlling for household income, and across all psychosocial outcomes, there was no interaction between gender and increased alcohol use, suggesting that the differences among genders in these psychosocial variables was the same between those who did and did not increase alcohol use since the start of the pandemic (Table 4). There was a significant interaction between increased alcohol use and pandemic phase (all *p* < .0001, Table 4). Pairwise tests indicated that at all phases, with the exception of pre-COVID, those who reported increased alcohol use had significantly more psychosocial symptoms on all measures (*p* < .0001, Table 4).

**Table 4.**
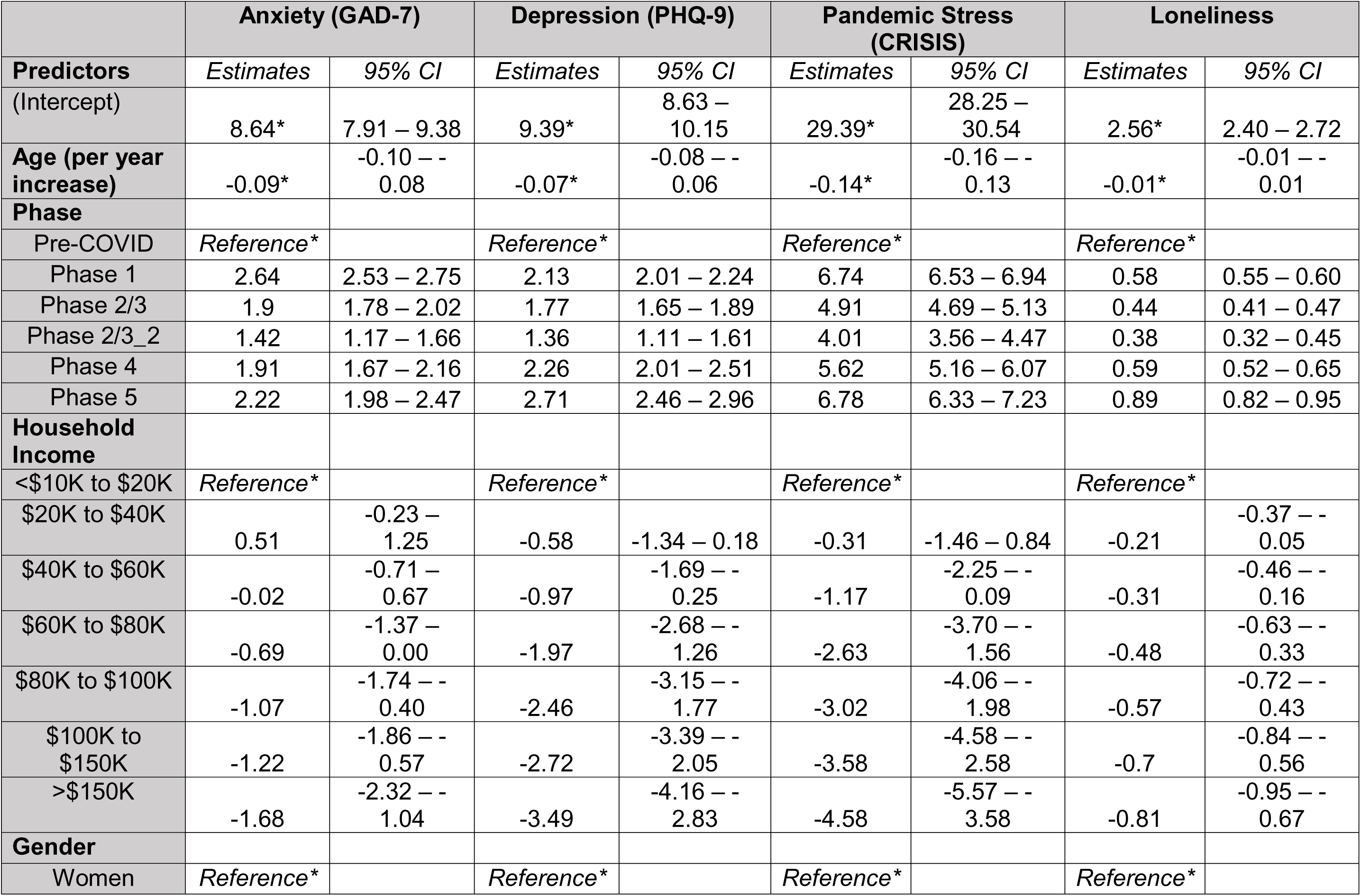
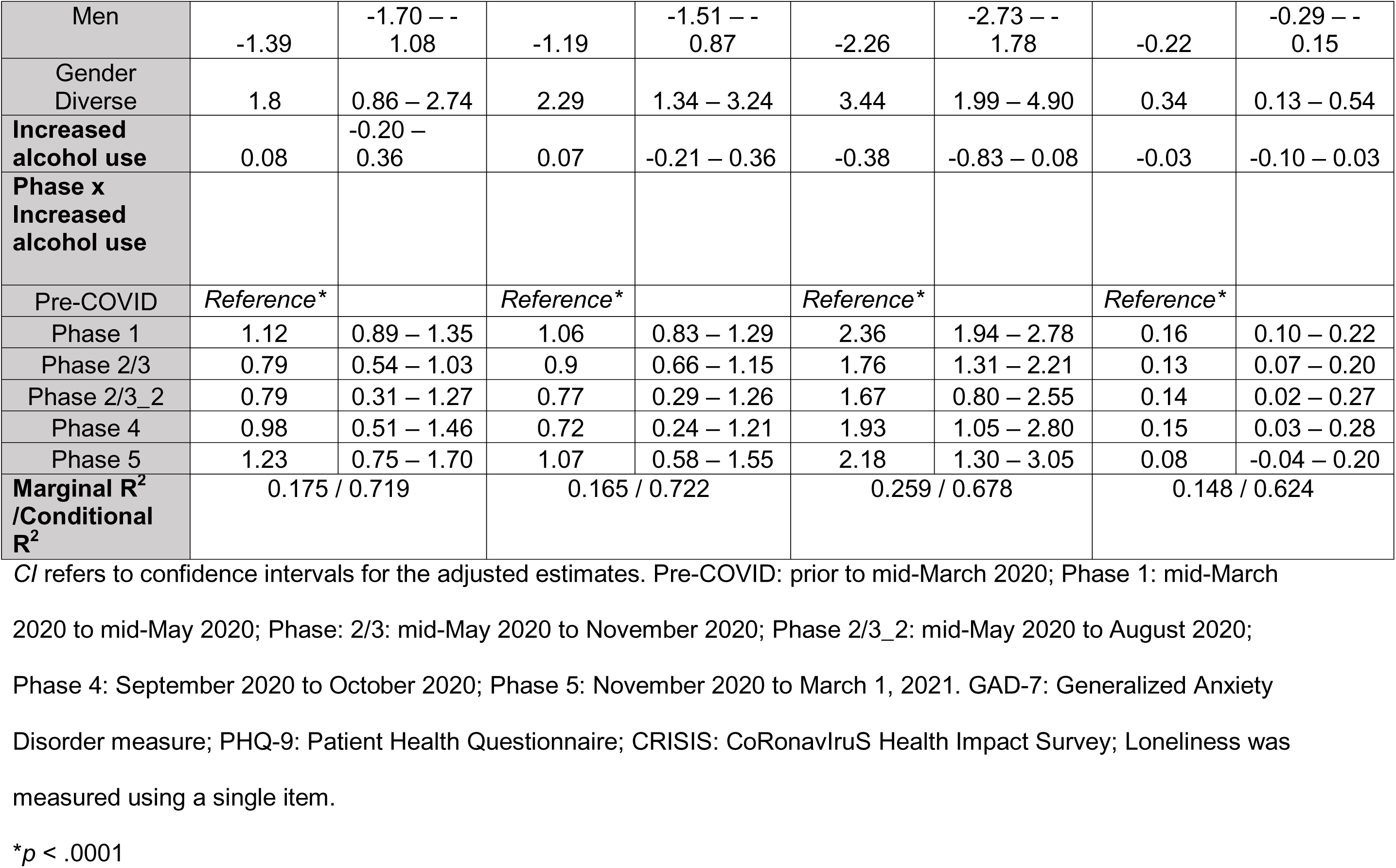
Psychosocial outcomes and sociodemographic factors for those that reported an increase in alcohol use.

### Associations between Psychosocial Outcomes and Cannabis Use by Gender

A total of 5.9% of the sample reported an increase in cannabis use since the start of the pandemic. Increased cannabis use was negatively associated with age (*p* <.001, Table 3), with older participants having lower odds of increased use. There was a significant relationship with gender (*p* = .02, Table 3, Fig 3), where women had a significantly higher odds of increased cannabis use compared to men, and there was no significant difference between men and gender diverse, and women and gender diverse groups .

**Fig 3.**
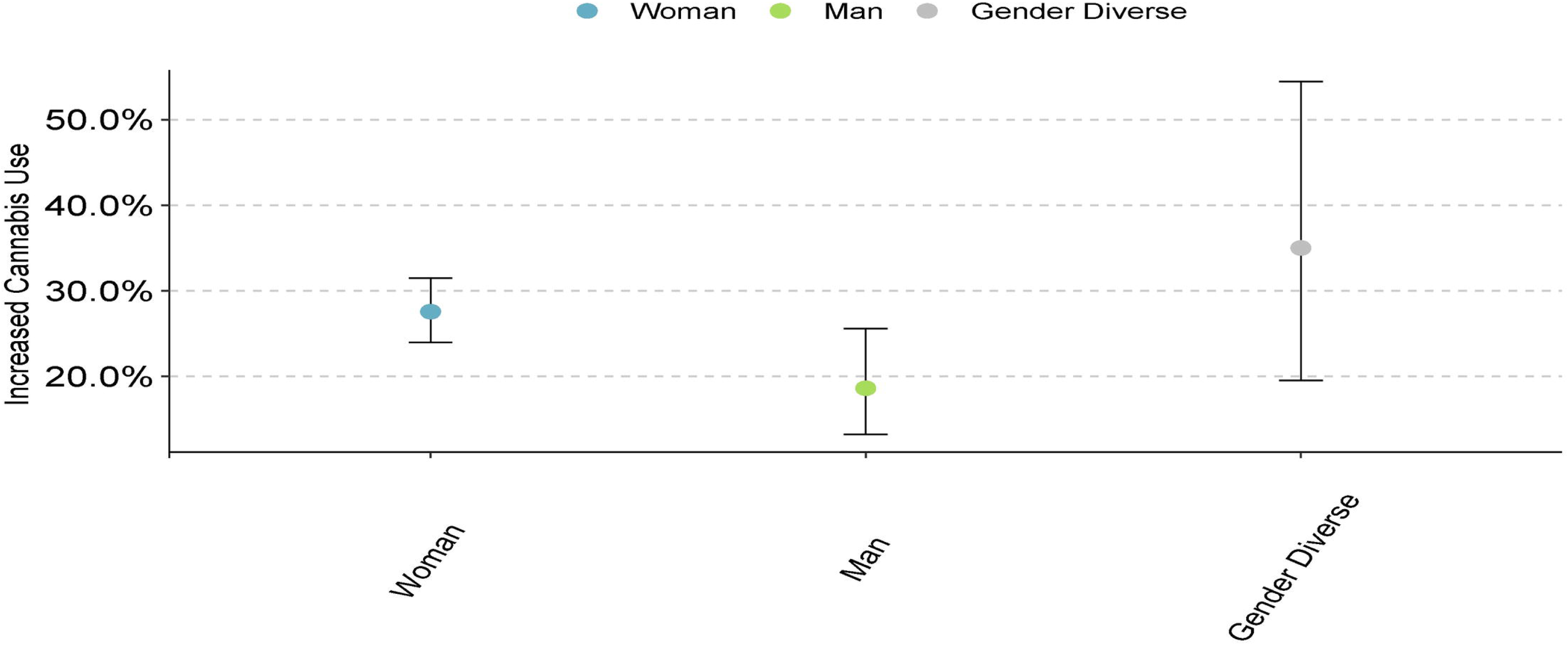
Cannabis use across different genders.

Controlling for household income, there was a significant interaction between change in cannabis use and pandemic phase (*p* < .0001 for GAD-7, PHQ-9, and CRISIS, *p* = .04 for Loneliness, Table 5). Post-hoc pairwise tests suggest that across all phases, including pre-COVID, those who increased cannabis use had significantly higher anxiety, more depressive symptoms, and higher COVID-stress scores than those who did not have increased cannabis use. Loneliness scores were significantly higher across all phases of the pandemic for those who increased cannabis use compared to those who did not. There was no interaction between gender and increased cannabis use for GAD-7, PHQ-9, or CRISIS scores. However, there was a significant interaction between gender and increased cannabis use on Loneliness (*p* = .008, Table 5). For both men and women, those who increased cannabis use had higher loneliness scores than those who did not have increased cannabis use. Conversely, among the gender-diverse participants, there was no difference in loneliness between those who increased cannabis, and those who did not.

**Table 5.**
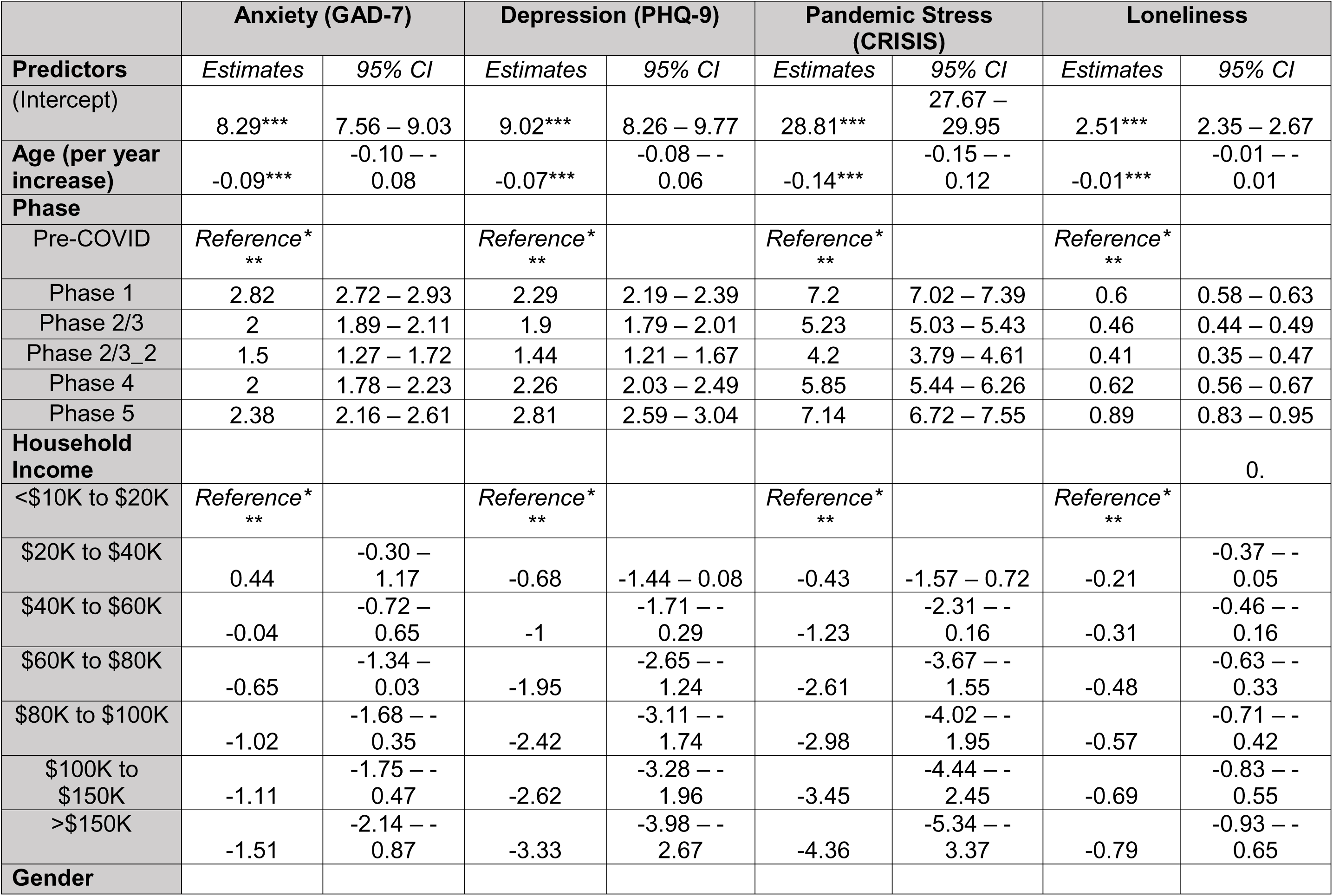
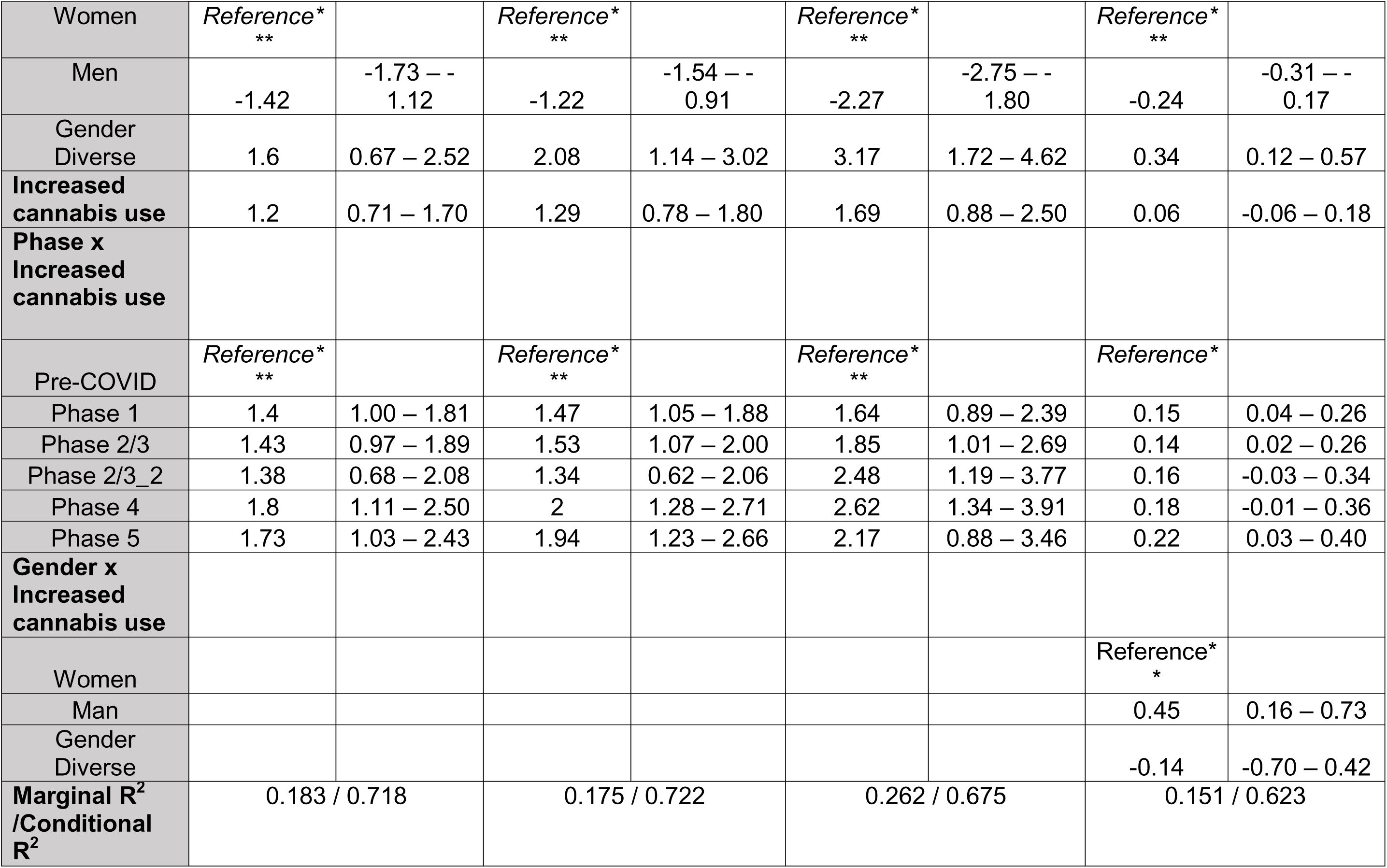
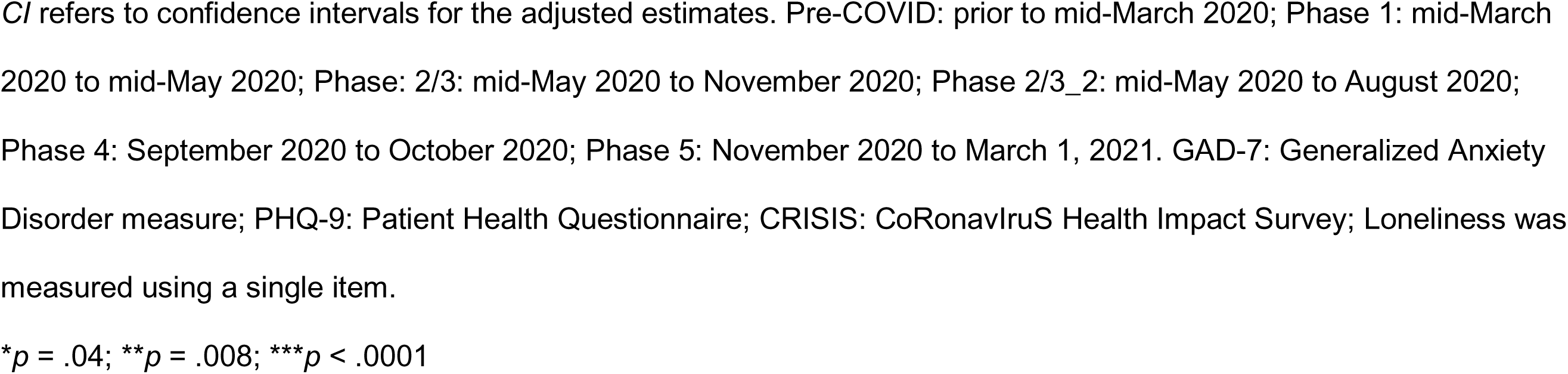
Psychosocial outcomes and sociodemographic factors for those that reported an increase in cannabis.

## Discussion

This large Canadian study recruited 6,076 women, men, and gender diverse people across the province of British Columbia. Our main findings indicated that age, sex, gender, ethnicity, Indigenous status, sexual orientation, and phase of the pandemic have distinct effects on psychosocial outcomes. Across outcomes, women had more psychosocial symptoms than men, regardless of their age or ethnicity, while the gender diverse group (n = 72) had even more symptoms than women. An analysis by sex revealed the same findings as for gender, except that the gender diverse group was now absorbed into one of the two binary categories and obscuring their findings.

Our results highlight the greater negative outcomes on all psychosocial variables in gender diverse individuals, which would have been obscured in an analysis by sex alone and adds to the literature highlighting the value in analyzing data by gender. It is important to underscore that being a woman was a significant factor that determined higher anxiety, depression, stress, and loneliness - a finding mirrored in the literature across all continents [38, 39]. Given that women and gender diverse individuals are more likely to be diagnosed with mood disorders or score lower on mood surveys outside of a pandemic [16,40,41], it is not surprising that these populations are experiencing mental health inequities during COVID-19. However, our results should be interpreted with some caution as our gender-diverse cohort accounted for only 1% of the sample. Nonetheless, our results are striking and consistent with many other studies focused on gender using larger cohorts [42, 43].

Our study also benefited from examining the effects of other intersectional sociodemographic variables, such as age, to determine how they might play a role in the effect of sex and gender on mental health. Across all the psychosocial measures, younger participants were more likely to have anxiety, depression, pandemic stress, and loneliness, irrespective of their gender. These findings are consistent with others in smaller cohort studies that indicated younger ages were associated with higher scores (i.e., more psychosocial symptoms) on psychosocial variables [44]. There may be several reasons for these findings such as restricted social engagements, barriers to employment, and living conditions. Lockdowns across the globe have resulted in restricted social gatherings, closing of restaurants, bars and clubs, as well as recreational sporting activities (gyms, sports clubs, exercise classes, yoga and dance). In addition, younger adults are more likely to either live on their own, or with unrelated roommates and have greater perceived lack of social support. Indeed, findings from a larger cohort in China found that greater loneliness was associated not only with younger age (16-29) but also in unmarried individuals [36]. Physical activity is another important factor as a large survey across fourteen countries found that decreased physical activity during restrictions and lockdowns, as well as high physical activity pre-pandemic, were associated with poorer mental health scores [45]. Other studies have also noted that suicide and suicidal ideation have increased during the pandemic in younger adults [46], related partially to job losses. Taken together, the underlying reasons for this significant effect of age are of great importance and require further study.

In addition to age, ethnicity was associated with psychosocial outcomes with Chinese/Taiwanese participants reporting significantly lower scores (i.e., fewer psychosocial symptoms) on anxiety, depression, pandemic stress, and loneliness. These data are consistent with findings from other studies, such as a survey of more than 46,000 Canadians which found that Chinese individuals were less likely to report symptoms consistent with moderate to severe generalized anxiety disorder than other visible minority groups during the COVID-19 pandemic [47]. Several studies of the Canadian Community Health Survey also show that the prevalence of mental disorders is lower among Asian or Chinese immigrants [48]. However, it is important that we interpret these findings with a consideration for how culture may affect the experience of mental well-being and distress. Culture has previously emerged as a major factor explaining differences in mental health consultation between Chinese and non-Chinese Canadian ethnic groups [48]. There is stigma in traditional Chinese culture around mental health problems and treatment, and prioritization of collective harmony over individual emotions [48]. The negative characterization of psychosocial experiences within Chinese cultural groups may affect disclosure of mental health issues and influence how individuals respond to questions about their mental health. For instance, when restricting analyses to only those at moderate to high risk for depression, Chinese Canadians were less likely to consult mental health services than non-Chinese Canadians in BC [48]. Similar findings have been demonstrated internationally [49]. In terms of the validity of these measures among Chinese/Taiwanese populations, the PHQ-9 has been validated for diagnosing major depressive disorder across major ethnic groups (African American, Chinese American, Latino, non-Hispanic White), suggesting that it can be used without adjustment in diverse populations [50]. On the other hand, while the feasibility, psychometric structure and construct validity of CRISIS, our measure of stress, was demonstrated in population-based assessments with broad coverage with respect to race, further work is required to assess CRISIS in different racial/ethnic groups [34]. Additionally, the GAD-7 was developed and validated among a primarily white sample, leaving open the possibility of a measurement bias [51], although it was concluded to be a reliable measure across some cultural groups [51, 52]. In sum, our findings suggest that our Chinese/Taiwanese sample experienced fewer psychosocial symptoms throughout the pandemic relative to other groups.

We found that those who self-identified as Indigenous had significantly worse psychosocial symptoms than non-Indigenous participants across all four scales for all phases of the pandemic in BC. Importantly, there was no difference in psychosocial outcomes between Indigenous and non-indigenous groups pre-COVID, which underscores the disproportionate impact of the pandemic on this community. While investigations on the mental health impacts on Indigenous peoples during the COVID-19 pandemic have been limited, our results are consistent with the available data. For example, other data from Australia (Aboriginal or Torres Strait Islander) [53] as well as Canada [54] showed more psychosocial symptoms among Indigenous respondents during the COVID-19 pandemic. The lack of interaction between Indigenous status and gender suggests that the higher psychosocial symptoms occur regardless of Indigenous persons’ gender. This stands in contrast to another study finding that Indigenous women were particularly impacted by mental health issues (severe generalized anxiety, worse mental health, and stress) during COVID-19 [54]. Future studies should explore the extent to which variables such as rurality (which can contribute to barriers accessing care) and income may account for these higher rates of psychological symptoms among Indigenous communities [44].

Findings on the relationship between anxiety, depression, pandemic stress, and loneliness, with increased alcohol and cannabis use, align with previous studies [12]. Given the poorer self-reported mental health among younger populations, it was not surprising to observe an increase in alcohol and cannabis use among this group, which suggests that alcohol use may be a form of coping for younger persons. We cannot attribute directionality to this association, nor eliminate the possibility that increased alcohol and cannabis use may be contributing to the increased psychosocial symptoms observed among younger populations during the pandemic. The lack of a gender difference in alcohol use increases contrasts with a previous American study [55] which found that females had increased their alcohol use compared to males. In contrast, we saw a gender effect on increased cannabis use, which was expressed by women, but not by men or gender diverse persons. In recent surveys, 28% of British Columbians had engaged in cannabis use in the past twelve months, compared to the Canadian average of 11%, suggesting that British Columbians are more likely to engage in cannabis use, and therefore may be more likely to use cannabis as a form of coping [56, 57]. Although cannabis use has been associated with male typicality and may go against gender norms typical to women [58], it may be that the social isolation and a lack of gatherings erased these social norms and facilitated women’s more active engagement in additional cannabis use, relative to pre-pandemic levels.

As predicted, psychosocial symptoms worsened over the course of the pandemic, with some of the worst scores observed early on, aligning with previous studies that found a higher prevalence of mental health disorders during the initial COVID-19 lockdown in March 2020 [59, 60]. Phase 2 and 3 of COVID restrictions in BC were characterized by an easing of restrictions, permitting outdoor gatherings and small social events, and the summer season. This loosening of public health measures was associated with a slight improvement in mental health, more than likely due to an increase in perceived social support and optimism regarding the state of the pandemic. Mental health outcomes were worsened in Phases 4 and 5 as BC entered wave 2 of the pandemic and public health orders tightened once again. It is important to note the average PHQ-9 and GAD-7 scores did not meet the criteria for clinical depression or anxiety, but that these levels increased relative to pre-pandemic levels as well as over time.

### Strengths and Limitations

Our study benefitted from a large, population-based sample size, and, despite known mental health disparities by gender, as far as we are aware, was one of the few that sought to explore findings from a gender lens by including gender-diverse groups as well, given known mental health disparities by gender [17]. That said, our sample size for gender diverse individuals was still limited [17]. Future studies should further investigate mental health in the gender diverse community during the COVID-19 pandemic with a focus on people of all ages, in contrast to previous studies [22]. Another limitation of the present study was the retrospective, cross-sectional nature of the survey, where participants completed the survey at only one time point, and were asked to retrospectively recall their mood and anxiety during different time points. This may have increased the likelihood of recall bias and reducing our capacity to examine causality and directionality of poor mental health outcomes. Finally, this study was confined to the general population of BC and therefore results may only be generalizable to the Canadian population, and populations with similar demographics to the present study.

### Implications

Our study has important implications for public health policy. These findings illustrate that government policies and interventions for future pandemics should place on emphasis on young adults, low-income populations, women, Indigenous, and gender diverse communities. Additionally, our study was one of the first to measure mental health outcomes across different phases of the pandemic, directly examining the effect of increased public health measures on mental health. At the time of writing, the vaccine rollout is well underway in BC with experts predicting an end to the pandemic in the months ahead, however, it is unclear whether mental health will return to pre-pandemic levels, or when life will return to “normal.” Moving forward, policy makers and leaders need to consider our findings when planning future public health measures. In future pandemics, the mental health of marginalized populations needs to be considered proactively. As vaccination efforts continue and case counts fall, it will also be critical to monitor the health status of these populations to ensure that they are not left behind. Additionally, for future pandemics and outbreaks, mobilizing resources to these communities early on can aid in mitigating these inequities from the beginning, rather than as an afterthought.

## Data Availability

All data in the manuscript is available and contained with the documents submitted. There are no additional links.

## Acknowledgements

We wish to thank Falla Jin and Shanlea Gordon (both at the BC Children’s Research Institute) for their assistance with data collection.

## Supplemental Information

**S1 Table. Public health measures during different phases of the COVID-19 pandemic in British Columbia.** Measures listed are not exhaustive.

## References

1. W.H.O. Weekly Operational Update on COVID-19 [Internet]. 2021 May. Available from: https://www.who.int/docs/default-source/coronaviruse/weekly-updates/wou_2021_3may_cleared-.pdf?sfvrsn=508fa040_3&download=true

2. PHAC. Canada COVID-19 Weekly Epidemiology Report [Internet]. 2021 May. Available from: https://www.canada.ca/content/dam/phac-aspc/documents/services/diseases/2019-novel-coronavirus-infection/surv-covid19-weekly-epi-update-20210514-eng.pdf

3. Wu P, Fang Y, Guan Z, Fan B, Kong J, Yao Z, et al. The Psychological Impact of the SARS Epidemic on Hospital Employees in China: Exposure, Risk Perception, and Altruistic Acceptance of Risk. Can J Psychiatry. 2008;54(5):302–11.

4. Wang Y, Xu B, Zhao G, Cao R, He X, Fu S. Is quarantine related to immediate negative psychological consequences during the 2009 H1N1 epidemic? Gen Hosp Psychiat. 2011;33(1):75–7.

5. Jalloh MF, Li W, Bunnell RE, Ethier KA, O’Leary A, Hageman KM, et al. Impact of Ebola experiences and risk perceptions on mental health in Sierra Leone, July 2015. Bmj Global Heal. 2018;3(2):e000471.

6. Wu T, Jia X, Shi H, Niu J, Yin X, Xie J, et al. Prevalence of mental health problems during the COVID-19 pandemic: A systematic review and meta-analysis. J Affect Disorders. 2020;281:91–8.

7. Cénat JM, Blais-Rochette C, Kokou-Kpolou CK, Noorishad P-G, Mukunzi JN, McIntee S-E, et al. Prevalence of Symptoms of Depression, Anxiety, Insomnia, Posttraumatic Stress Disorder, and Psychological Distress among Populations Affected by the COVID-19 Pandemic: A Systematic Review and Meta-Analysis. Psychiat Res. 2020;295:113599.

8. Liu D, Ren Y, Yan F, Li Y, Xu X, Yu X, et al. Psychological Impact and Predisposing Factors of the Coronavirus Disease 2019 (COVID-19) Pandemic on General Public in China. Ssrn Electron J. 2020;

9. Wong SYS, Zhang D, Sit RWS, Yip BHK, Chung RY, Wong CKM, et al. Impact of COVID-19 on loneliness, mental health, and health service utilisation: a prospective cohort study of older adults with multimorbidity in primary care. Brit J Gen Pract. 2020;70(700):bjgp20X713021.

10. Stanton R, To QG, Khalesi S, Williams SL, Alley SJ, Thwaite TL, et al. Depression, Anxiety and Stress during COVID-19: Associations with Changes in Physical Activity, Sleep, Tobacco and Alcohol Use in Australian Adults. Int J Environ Res Pu. 2020;17(11):4065.

11. Vanderbruggen N, Matthys F, Laere SV, Zeeuws D, Santermans L, Ameele SV den, et al. Self-Reported Alcohol, Tobacco, and Cannabis Use during COVID-19 Lockdown Measures: Results from a Web-Based Survey. Eur Addict Res. 2020;26(6):309–15.

12. Dumas TM, Ellis W, Litt DM. What Does Adolescent Substance Use Look Like During the COVID-19 Pandemic? Examining Changes in Frequency, Social Contexts, and Pandemic-Related Predictors. J Adolescent Health. 2020;67(3):354–61.

13. Wang Y, Kala MP, Jafar TH. Factors associated with psychological distress during the coronavirus disease 2019 (COVID-19) pandemic on the predominantly general population: A systematic review and meta-analysis. Plos One. 2020;15(12):e0244630.

14. Pieh C, Budimir S, Probst T. The effect of age, gender, income, work, and physical activity on mental health during coronavirus disease (COVID-19) lockdown in Austria. J Psychosom Res. 2020;136:110186.

15. Hawkes S, Tanaka S, Pantazis A, Gautam A, Kiwuwa-Muyingo S, Buse K, et al. Recorded but not revealed: exploring the relationship between sex and gender, country income level, and COVID-19. Lancet Global Heal. 2021;9(6):e751–2.

16. Eid RS, Gobinath AR, Galea LAM. Sex Differences in Depression: Insights from Clinical and Preclinical Studies. Prog Neurobiol. 2019;176:86–102.

17. Jiskrova GK, Bobák M, Pikhart H, Ksinan AJ. Job loss and lower healthcare utilisation due to COVID-19 among older adults across 27 European countries. J Epidemiol Commun H. 2021;jech-2021-216715.

18. Özdin S, Özdin ŞB. Levels and predictors of anxiety, depression and health anxiety during COVID-19 pandemic in Turkish society: The importance of gender. Int J Soc Psychiatr. 2020;66(5):504–11.

19. Smith L, Jacob L, Yakkundi A, McDermott D, Armstrong NC, Barnett Y, et al. Correlates of symptoms of anxiety and depression and mental wellbeing associated with COVID-19: a cross-sectional study of UK-based respondents. Psychiat Res. 2020;291:113138.

20. Qiu J, Shen B, Zhao M, Wang Z, Xie B, Xu Y. A nationwide survey of psychological distress among Chinese people in the COVID-19 epidemic: implications and policy recommendations. Gen Psychiatry. 2020;33(2):e100213.

21. Gobinath AR, Mahmoud R, Galea LAM. Influence of sex and stress exposure across the lifespan on endophenotypes of depression: focus on behavior, glucocorticoids, and hippocampus. Front Neurosci-switz. 2015;8:420.

22. Hawke LD, Hayes E, Darnay K, Henderson J. Mental health among transgender and gender diverse youth: An exploration of effects during the COVID-19 pandemic. Psychology Sex Orientat Gend Divers. 2021;

23. TransPULSECanada. Health and health care access for trans and non-binary people in Canada [Internet]. 2020 Mar. Available from: https://transpulsecanada.ca/research-type/reports

24. Braveman P, Gottlieb L. The Social Determinants of Health: It’s Time to Consider the Causes of the Causes. Public Health Rep. 2014;129(1_suppl2):19–31.

25. Hawkins D. Social Determinants of COVID-19 in Massachusetts, United States: An Ecological Study. J Prev Medicine Public Heal. 2020;53(4):220–7.

26. Sun S, Pachankis JE, Li X, Operario D. Addressing Minority Stress and Mental Health among Men Who Have Sex with Men (MSM) in China. Curr Hiv-aids Rep. 2020;17(1):35–62.

27. Fiorillo A, Sampogna G, Giallonardo V, Vecchio VD, Luciano M, Albert U, et al. Effects of the lockdown on the mental health of the general population during the COVID-19 pandemic in Italy: Results from the COMET collaborative network. Eur Psychiat. 2020;63(1):1–28.

28. Brotto L. Early Release: COVID-19 Rapid Evidence Study of a Provincial Population-Based Cohort for Gender and Sex (RESPPONSE): Methods [Internet]. 2021 Mar. Available from: https://whri.org/wp-content/uploads/2021/03/COVID-19-RESPPONSE_Methods-Report_March-11-2021_FINAL.pdf

29. Harris PA, Taylor R, Thielke R, Payne J, Gonzalez N, Conde JG. Research electronic data capture (REDCap)—A metadata-driven methodology and workflow process for providing translational research informatics support. J Biomed Inform. 2009;42(2):377–81.

30. Kroenke K, Spitzer RL, Williams JBW. The PHQ-9. J Gen Intern Med. 2001;16(9):606–13.

31. Huang FY, Chung H, Kroenke K, Delucchi KL, Spitzer RL. Using the patient health questionnaire-9 to measure depression among racially and ethnically diverse primary care patients. J Gen Intern Med. 2006;21(6):547–52.

32. Spitzer RL, Kroenke K, Williams JBW, Löwe B. A Brief Measure for Assessing Generalized Anxiety Disorder: The GAD-7. Arch Intern Med. 2006;166(10):1092–7.

33. Löwe B, Decker O, Müller S, Brähler E, Schellberg D, Herzog W, et al. Validation and Standardization of the Generalized Anxiety Disorder Screener (GAD-7) in the General Population. Med Care. 2008;46(3):266–74.

34. Nikolaidis A, Paksarian D, Alexander L, Derosa J, Dunn J, Nielson DM, et al. The Coronavirus Health and Impact Survey (CRISIS) reveals reproducible correlates of pandemic-related mood states across the Atlantic. Sci Rep-uk. 2021;11(1):8139.

35. Palgi Y, Shrira A, Ring L, Bodner E, Avidor S, Bergman Y, et al. The Loneliness Pandemic: Loneliness and Other Concomitants of Depression, Anxiety and their Comorbidity During the COVID-19 Outbreak. J Affect Disorders. 2020;275:109–11.

36. Stravynski A, Boyer R. Loneliness in Relation to Suicide Ideation and Parasuicide: A Population-Wide Study. Suicide Life-threat. 2001;31(1):32–40.

37. McClelland H, Evans JJ, Nowland R, Ferguson E, O’Connor RC. Loneliness as a predictor of suicidal ideation and behaviour: a systematic review and meta-analysis of prospective studies. J Affect Disorders. 2020;274:880–96.

38. Gasteiger N, Vedhara K, Massey A, Jia R, Ayling K, Chalder T, et al. Depression, anxiety and stress during the COVID-19 pandemic: results from a New Zealand cohort study on mental well-being. Bmj Open. 2021;11(5):e045325.

39. Mautong H, Gallardo-Rumbea JA, Alvarado-Villa GE, Fernández-Cadena JC, Andrade-Molina D, Orellana-Román CE, et al. Assessment of depression, anxiety and stress levels in the Ecuadorian general population during social isolation due to the COVID-19 outbreak: a cross-sectional study. Bmc Psychiatry. 2021;21(1):212.

40. Gutiérrez-Lobos K, Scherer M, Anderer P, Katschnig H. The influence of age on the female/male ratio of treated incidence rates in depression. Bmc Psychiatry. 2002;2(1):3.

41. Stanton AM, Batchelder AW, Kirakosian N, Scholl J, King D, Grasso C, et al. Differences in mental health symptom severity and care engagement among transgender and gender diverse individuals: Findings from a large community health center. Plos One. 2021;16(1):e0245872.

42. Jarrett BA, Peitzmeier SM, Restar A, Adamson T, Howell S, Baral S, et al. Gender-affirming care, mental health, and economic stability in the time of COVID-19: a global cross-sectional study of transgender and non-binary people. Medrxiv. 2020;2020.11.02.20224709.

43. Restar AJ, Jin H, Jarrett B, Adamson T, Baral SD, Howell S, et al. Characterising the impact of COVID-19 environment on mental health, gender affirming services and socioeconomic loss in a global sample of transgender and non-binary people: a structural equation modelling. Bmj Global Heal. 2021;6(3):e004424.

44. Bao L, Li W-T, Zhong B-L. Feelings of loneliness and mental health needs and services utilization among Chinese residents during the COVID-19 epidemic. Globalization Health. 2021;17(1):51.

45. Wilke J, Hollander K, Mohr L, Edouard P, Fossati C, González-Gross M, et al. Drastic Reductions in Mental Well-Being Observed Globally During the COVID-19 Pandemic: Results From the ASAP Survey. Frontiers Medicine. 2021;8:578959.

46. Ueda M, Nordström R, Matsubayashi T. Suicide and mental health during the COVID-19 pandemic in Japan. J Public Health. 2021;

47. Moyser M. The mental health of population groups designated as visible minorities in Canada during the COVID-19 pandemic [Internet]. 2020 Sep. Available from: https://www150.statcan.gc.ca/n1/en/pub/45-28-0001/2020001/article/00077-eng.pdf?st=GJKdbqci

48. Chen AW, Kazanjian A, Wong H. Why do Chinese Canadians not Consult Mental Health Services: Health Status, Language or Culture? Transcult Psychiatry. 2009;46(4):623–41.

49. Abe-Kim J, Takeuchi DT, Hong S, Zane N, Sue S, Spencer MS, et al. Use of Mental Health–Related Services Among Immigrant and US-Born Asian Americans: Results From the National Latino and Asian American Study. Am J Public Health. 2007;97(1):91–8.

50. Huang Y, Zhao N. Mental health burden for the public affected by the COVID-19 outbreak in China: Who will be the high-risk group? Psychology Heal Medicine. 2020;26(1):1–12.

51. Parkerson HA, Thibodeau MA, Brandt CP, Zvolensky MJ, Asmundson GJG. Cultural-based biases of the GAD-7. J Anxiety Disord. 2015;31:38–42.

52. Zhou Y, Xu J, Rief W. Are comparisons of mental disorders between Chinese and German students possible? An examination of measurement invariance for the PHQ-15, PHQ-9 and GAD-7. Bmc Psychiatry. 2020;20(1):480.

53. Newby JM, O’Moore K, Tang S, Christensen H, Faasse K. Acute mental health responses during the COVID-19 pandemic in Australia. Plos One. 2020;15(7):e0236562.

54. Arriagada P, O’Donnell TH and V, O’Donnell V. Indigenous people and mental health during the COVID-19 pandemic [Internet]. 2020 Jun. Available from: https://www150.statcan.gc.ca/n1/en/pub/45-28-0001/2020001/article/00035-eng.pdf?st=eJZ0mZ3X

55. Rodriguez LM, Litt DM, Stewart SH. Drinking to cope with the pandemic: The unique associations of COVID-19-related perceived threat and psychological distress to drinking behaviors in American men and women. Addict Behav. 2020;110:106532.

56. BCStats. Cannabis in British Columbia: Results from the 2018 BC Cannabis Use Survey [Internet]. 2019 Jun. Available from: https://www2.gov.bc.ca/assets/gov/data/statistics/health-safety/cannabis_bc_results_2018_survey.pdf

57. GoC. Canadian Cannabis Survey 2020: Summary [Internet]. 2020 [cited 2021 May 16]. Available from: https://www.canada.ca/en/health-canada/services/drugs-medication/cannabis/research-data/canadian-cannabis-survey-2020-summary.html

58. Hemsing N, Greaves L. Gender Norms, Roles and Relations and Cannabis-Use Patterns: A Scoping Review. Int J Environ Res Pu. 2020;17(3):947.

59. Gualano MR, Moro GL, Voglino G, Bert F, Siliquini R. Effects of Covid-19 Lockdown on Mental Health and Sleep Disturbances in Italy. Int J Environ Res Pu. 2020;17(13):4779.

60. Niedzwiedz CL, Green MJ, Benzeval M, Campbell D, Craig P, Demou E, et al. Mental health and health behaviours before and during the initial phase of the COVID-19 lockdown: longitudinal analyses of the UK Household Longitudinal Study. J Epidemiol Commun H. 2021;75(3):224–31.

