## Supplemental Table 1 for "The influence of sex, gender, age, and ethnicity on psychosocial factors and substance use throughout phases of the COVID-19 pandemic"

| **COVID-19 Phases** | **Public Health Control Measures in British Columbia** |
| --- | --- |
| Phase 1 (mid-March 2020 to mid-May 2020) | Closing of all businesses and a ban on gatherings, while essential services remain open. Essential services include, but are not limited to: essential health services, transportation, food and agriculture service providers, liquor and cannabis stores, and vulnerable population service providers. |
| Phase 2 (mid-May 2020 to mid-June 2020) | The start of reopening in British Columbia, including hair salons, restaurants, libraries, office-based worksites, sports and childcare. Students K-12 returned to school on a gradual and part-time basis. |
| Phase 3 (mid-June 2020 to August 2020) | A continued reopening including non-essential travel within the province, the re-opening of the accommodation industry, movie theatres. |
| Phase 4 (September 2020 to October 2020) | Restrictions tighten once again including a 10PM last call for liquor service, prohibition of events in banquet halls, and a cap on social gatherings |
| Phase 5 (November 2020 to March 1, 2021) | Further restrictions placed on gatherings and services including a ban on gatherings at private residences for those outside of that household, mandatory masks in indoor and crowded settings, restrictions of sports facilities and gyms |
